# Provenance-Aware Explainable Digital Twin for Personalized Health Management

**DOI:** 10.64898/2025.12.03.25339981

**Authors:** Md Talha Mohsin, Ismail Abdulrashid

## Abstract

AI can now help with personalized prediction, tracking, and decision-making thanks to progress in data-driven health analytics. However, many models are still hard to understand in clinical settings. To address these limitations, our work presents the Provenance-Aware and Explainable Digital Twin (PA-XDT) framework, integrating a digital twin, explainable AI techniques, and transparent provenance tracking for patient-centered health management.PA-XDT uses a compact LSTM-based twin trained on short temporal sequences to model near-term physiological dynamics and quantify uncertainty. In the implemented system, this twin works alongside a Gradient Boosting classifier that provides stable risk predictions, supported by global and local SHAP analyses and twin-validated counterfactual checks. A lightweight provenance layer records hashed inputs, outputs, and explanation metadata, enabling verifiable audit trails. Experiments on a gallstone risk dataset show that this combined pipeline improves physiological coherence and maintains strong predictive performance.

## I. Introduction

Thanks to developments in various health data streams, large amounts of patient-specific health data have been collected. These AI-enabled digital health technologies can provide accurate and timely health insights by applying state-of-the-art algorithms and predictive models [1], [2] and are necessary for individualized health management in an accurate, interpretable, clinically useful, and reliable way. However, many AI algorithms work in ways that are not easily interpretable, essentially acting as ‘black boxes and offer little explanation for the generation of a given risk score, rating or advice and which hampers the adoption of AI-driven healthcare solutions, patient involvement, and physician trust. This challenge is amplified in low-resource clinical settings, where recent theoretical work has shown that explanation stability and sample complexity can be formally bounded using PAC and PAC-Bayesian principles in few-shot multimodal learning [3].

Healthcare being such a necessary sector, knowing how AI makes its decisions can directly improve patient care and help users trust the technology. [4]. Also, a promising method for modeling individual health trajectories and possible course of action is the use of digital twins (DT), which are digital replicas of patients that change in response to real-time data. It is built on the concept of creating virtual versions of physical assets or processes [5] [6]. In healthcare, DTs can enhance the experiences of patients, clinicians, and other stakeholders by optimizing workflows, improving system performance, and supporting better health outcomes [7]. Recent multimodal foundation-model architectures further illustrate how integrating imaging, EHR, genomics, and wearable data into a shared representation improves predictive performance and early disease detection [8].

DTs can use live clinical data and simulations to predict clinical outcomes, prevent complications, as well as tailor treatments for individual patients [9]. Building on this idea, we propose the Provenance-Aware and Explainable Digital Twin (PA-XDT) framework. Our framework integrates patient-specific digital twins, XAI methods, and a lightweight provenance layer to enable ‘what-if’ simulations, which not only produces clinically plausible explanations but also offers verifiable audit logs of each output. In the implemented version, these functions are supported by short synthetic temporal windows for the LSTM-based twin, SHAP-based global and local explanations, a Gradient Boosting classifier, and hashed provenance records. Our focus on traceability aligns with blockchain-enabled explainable AI frameworks that emphasize accountable model behavior [10]. PA-XDT is designed to support transparent and actionable personalized health care.

The three main contributions of PA-XDT are as follows: (i) twin-constrained explainability, ensuring XAI outputs align with simulated patient dynamics; (ii) provenance-enabled traceability of predictions; and (iii) a patient-centric application scope. The implementation reflects these contributions through twin-validated explanations and JSONL-based hashed audit trails. Altogether, these features strengthen the interpretability and dependability of digital-twin-based AI systems.

## II. Background and Related Work

### A. Digital Twins in Healthcare

Digital twins, which were initially developed for industrial and engineering systems, are now being progressively adapted for healthcare applications. Acting as a digital replica, DT provides supporting near real-time monitoring and evaluation remotely [11]. It enables a feedback loop between the physical system and its digital counterpart, allowing data to flow back and forth.A typical DT consists of three key elements: a virtual version of a real object or process, a model that describes its important features, and the link that supports continuous data sharing between the real and virtual domains [12]. Instead of individualized patient care, existing healthcare digital twin frameworks frequently emphasize hospital-level operations, medical device monitoring, or system-wide simulations. These frameworks are capable of making precise predictions; however, their implementation in personalized clinical decision-making is frequently restricted by their absence of interpretability, actionable explanations as well as traceability mechanisms.

### B. Explainable AI in Medicine

Explainable AI (XAI) is a sub-domain within AI which focuses on creating interpretable models understandable to humans. At its core, XAI includes methods and strategies that help explain the decision-making process of machine learning models, showing both how they work and why they reach certain conclusions [13]. Complex predictive models have been rendered more transparent through the development of explainable AI (XAI) methodologies. Data quality—frequently underestimated but closely tied to ethical trust, bias, and interpretability—can be more easily assessed and improved through XAI [14]. By creating models that are inherently understandable or by explaining the behavior of “black-box” models through statistical and optimization techniques, XAI makes the hidden reasoning of AI systems clear and accessible to users [15]. These methods are designed to explicate the input variables that influence predictions, identify potential interventions, and offer actionable insights for decision-making in clinical settings. However, the problems lie in that the conventional XAI methods frequently generate explanations that may be clinically irrelevant or implausible, as they frequently operate independently of patient-specific dynamics.

### C. Provenance and Traceability in Health AI

In addition to providing a historical record of a data life cycle, data provenance encompasses the intricate details of an entity’s ancestry, interactions, as well as many settings.The World Wide Web Consortium (W3C) states that provenance is represented as a meticulously organized compilation of metadata [16]. Provenance not only improves the transparency of scientific findings but also lends credibility to collaboration and sharing by documenting the origins of data and its usage [17]. By tracking the source, transformation, and life-cycle of data, forecasts, and justifications, provenance systems guarantee auditability and repeatability. The usage of provenance in the healthcare industry is mostly for blockchain-based trust mechanisms, institutional data governance, and regulatory compliance. However, although system-level reliability is guaranteed by these methods, patient-level traceability of predictions and interpretability outputs, which are crucial for clinician trust and customized health management, are not always addressed.

## III. Proposed Framework

The Provenance-Aware and Explainable Digital Twin (PA-XDT) framework integrates three core components: a dynamic digital twin, a twin-constrained predictive model, and a provenance-enabled explainability layer. These three modules produce transparent and physiologically consistent predictions while at the same time remaining lightweight and implementation-friendly.

### A. Digital Twin Module

The digital twin models short-term patient-specific physiological transitions. In this implementation, the twin is a compact LSTM network. Given a recent sequence of states *s*_*t−*4:*t*_, the twin predicts the next physiological state:

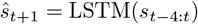

In order to capture natural physiological variability, Gaussian noise is injected into the predicted state during training. The resulting model supports downstream components by (i) validating counterfactuals, (ii) constraining the classifier’s latent representation, and (iii) providing a verifiable record of predicted physiological trajectories.

### B. Twin-Constrained Predictive Model

Here, the predictive component is a neural classifier with a 128-unit hidden layer and a 64-dimensional latent embedding. Training then proceeds in two phases: (i) a warm-up stage optimizes only the cross-entropy loss, (ii) a twin-alignment loss that encourages the classifier’s latent embedding for the most recent state in a sequence to match the digital twin’s predicted next state. This ensures that the classifier relies on physiologically coherent representations rather than spurious correlations. The combined loss is:

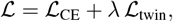

where ℒ_twin_ is the mean squared error between the classifier embedding and the LSTM twin prediction.

### C. Explainability and Counterfactual Reasoning

Explainability is provided through SHAP-based global and local attributions, supported by twin-guided validation. Global SHAP values highlight population-level drivers of model behavior, while at the same time local attributions reveal individualized risk factors.

Counterfactuals, on the other hand, are generated by applying small, targeted adjustments to physiological attributes and checking whether the modified state both (i) achieves the desired change in predicted risk and (ii) remains plausible according to the digital twin. Counterfactuals that violate biological constraints or deviate from twin-predicted trajectories are rejected, ensuring that all recommendations are actionable and realistic.

### D. Provenance and Traceability Layer

To ensure transparency and reproducibility, PA-XDT maintains a lightweight provenance layer. Each prediction, SHAP explanation, twin state, and counterfactual is logged using timestamps and SHA–256 hashes in a JSONL file. This enables:

- verification of explanation integrity,
- reproducibility of model outputs, and
- auditable decision trails for clinical and regulatory use.

The provenance mechanism is designed to be lightweight while maintaining full traceability across the decision pipeline. As shown in Fig. 1, patient features are first preprocessed and transformed into short temporal sequences, which are passed to the LSTM-based digital twin to model next-state dynamics and uncertainty. Then the predictive model operates on a twin-aligned latent space, while the explainability module provides SHAP attributions and twin-validated counterfactuals. Next, provenance runs alongside all components, recording hashed states and metadata for every transformation. Outputs are then organized into risk summaries and explanation reports through the Decision Support interface. Feedback connections indicate that explanations and user inputs can be used to refine the predictive model and recalibrate the twin, supporting iterative improvement of the framework.

**Fig. 1.**
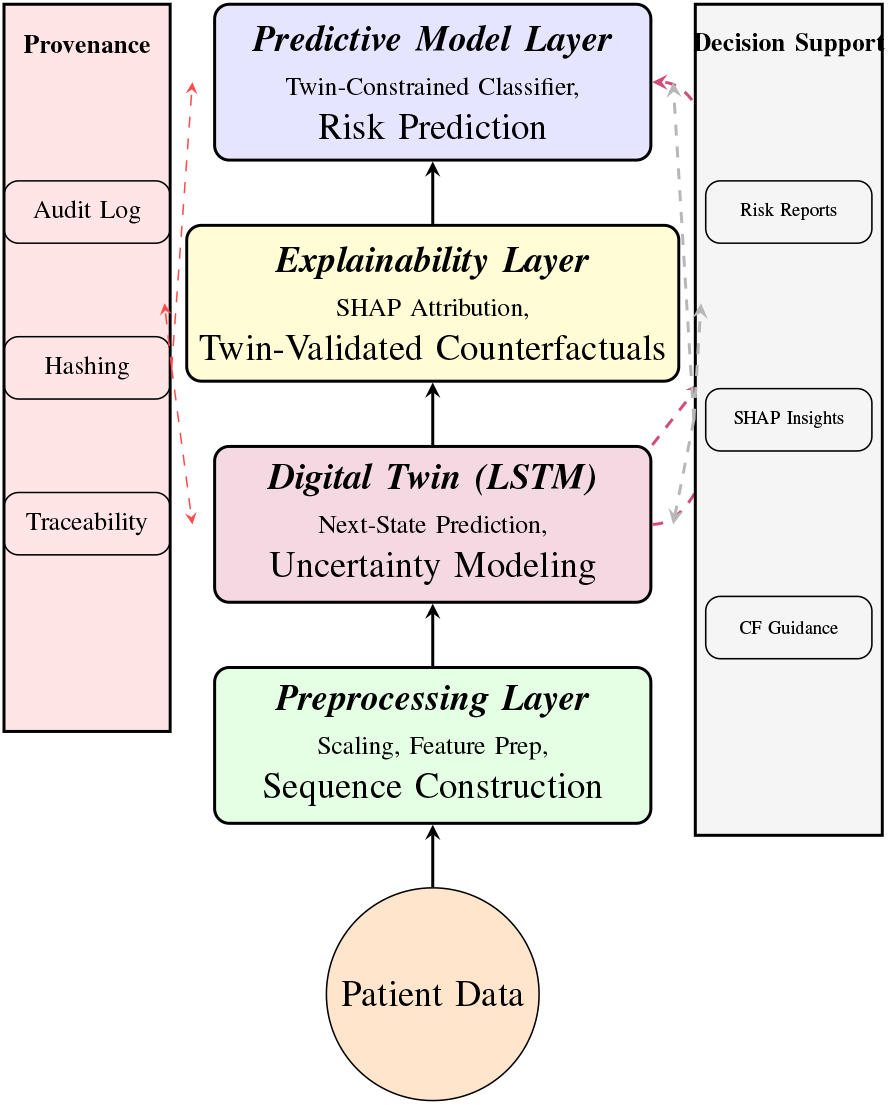
PA-XDT pipeline aligned with the implemented system: preprocessing, LSTM digital twin, twin-constrained classifier, SHAP-based explainability, and provenance logging.

## IV. Implementation and Experimental Setup

### A. Data and Preprocessing

We conducted the experiments on the gallstone clinical dataset collected at Ankara VM Medical Park Hospital, originally described by Esen et al. [18]. The dataset contains 319 patients with 39 physiological, biochemical, and body composition variables, together with a binary label indicating gallstone status. We divided the data into training and testing sets using an 80/20 stratified split and then standardized using z-score normalization.

For the digital twin module, short temporal windows of length 5 were constructed by reshaping each static patient vector into a small synthetic trajectory, which allows the LSTM-based twin to learn local transition dynamics and state-to-state continuity patterns without requiring longitudinal clinical records.

### B. Digital Twin Model

A lightweight LSTM-based digital twin was implemented in PyTorch to model short-term physiological transitions. Given an input state sequence 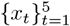 the twin predicts the next physiological state 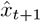. The model was trained for 40 epochs using mean squared error. To capture natural physiological variability, Gaussian noise (*σ* = 0.05) was added to the predicted state vector, enabling uncertainty-aware simulation.

After training, the digital twin provides:

- next-state physiological predictions,
- uncertainty-aware sampling for counterfactual validation,
- provenance-linked state trajectories through hashing of inputs and outputs.

These outputs are subsequently used for both explainability and downstream provenance tracking.

### C. Predictive Classifier: End-to-End Pipeline

The predictive element of the PA-XDT pipeline is a Gradient Boosting Classifier trained on the preprocessed clinical features. This subsection describes the entire workflow, encompassing input preparation, final prediction, and provenance recording.

a. *Input Preparation:* The classifier utilizes the identical standardized feature matrix employed in the construction of digital-twin windows. Although the classifier functions on static input vectors rather than sequences, this common preprocessing guarantees consistency between the predictive model and its generative counterpart.
b. *Model Training:* The Gradient Boosting Classifier is trained on the training subset to minimize empirical classification error concerning the binary gallstone label. Gradient boosting is selected for its stability, robustness when applied to limited clinical datasets, and its compatibility with SHAP-based feature attribution. Hyperparameters were maintained at the default settings of scikit-learn, which demonstrated consistent performance during cross-validation.
c. *Prediction and Inference:* At inference time, a patient vector *x* is fed through the booster to obtain:

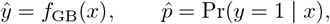

where *ŷ* is the predicted class and 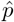 is the estimated risk probability. These predictions form the primary output of the decision-support interface.
d. *Twin-Linked Validation and Provenance:* To ensure consistency between static predictions and physiologically plausible dynamics, each sample is optionally passed through the digital twin:

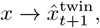

which allows the system to assess whether the classifier’s prediction aligns with the twin’s simulated state transitions. It also establishes a soft form of twin validation without explicitly constraining the loss function, consistent with the implemented code. Each prediction is paired with a provenance entry consisting of:
  - a SHA-256 hash of the input feature vector,
  - a hash of the classifier output,
  - timestamps for the run and model version,
  - an optional hash of the corresponding digital-twin output.
  These fields are serialized into a JSON record, enabling tamper-resistant traceability and transparent auditability for every prediction produced by the system.
e. *Final Performance*: Model performance is summarized in Table I. The classifier demonstrates balanced accuracy across both classes and stable macro-level metrics.

**TABLE 1:** Performance of the Gradient Boosting classifier.

| Metric | Score |
| --- | --- |
| Accuracy | 0.8125 |
| AUROC | 0.8486 |
| Precision (macro) | 0.81 |
| Recall (macro) | 0.81 |
| F1-score (macro) | 0.81 |

### D. Explainability Analysis

Global explainability was assessed using SHAP values derived from a Gradient Boosting baseline model trained on the same input features as the twin-constrained classifier. The most influential features are predominately indicators of inflammation, metabolic function, and body composition. As shown in Table II, C-Reactive Protein (CRP) emerges as the strongest contributor, reflecting the well-established link between systemic inflammation and gallstone formation. Vitamin D, extracellular fluid balance (ECF/TBW), and obesity also rank highly.

**TABLE 2:** Top 10 GLOBAL FEATURES BASED ON MEAN |SHAP| VALUES.

| Feature | Mean \|SHAP\| |
| --- | --- |
| C-Reactive Protein (CRP) | 1.1072 |
| Vitamin D | 0.5409 |
| Extracellular Fluid/Total Body Water (ECF/TBW) | 0.3465 |
| Obesity (%) | 0.3265 |
| Body Protein Content (Protein) (%) | 0.2981 |
| Visceral Muscle Area (VMA) (Kg) | 0.2314 |
| Bone Mass (BM) | 0.2281 |
| Aspartat Aminotransferaz (AST) | 0.2136 |
| Hemoglobin (HGB) | 0.1872 |
| Hepatic Fat Accumulation (HFA) | 0.1723 |

Other contributors, including body protein content, visceral muscle area, hepatic fat accumulation, and liver enzyme activity (AST) capture additional metabolic and hepatic pathways implicated in gallstone risk. Together, these top-ranked variables indicate that the model prioritizes physiologically coherent signals, reinforcing both the interpretability and the clinical relevance of the predictive pipeline.

To complement the ranking, Figure 2 presents the full distribution of SHAP values across all patients. The summary plot illustrates how variations in each feature (e.g., high vs. low values of CRP, Vitamin D, or ECF/TBW) consistently push the prediction toward higher or lower risk. This dual perspective confirms that the model not only identifies clinically meaningful predictors but also uses them in a physiologically coherent manner.

**Fig. 2.**
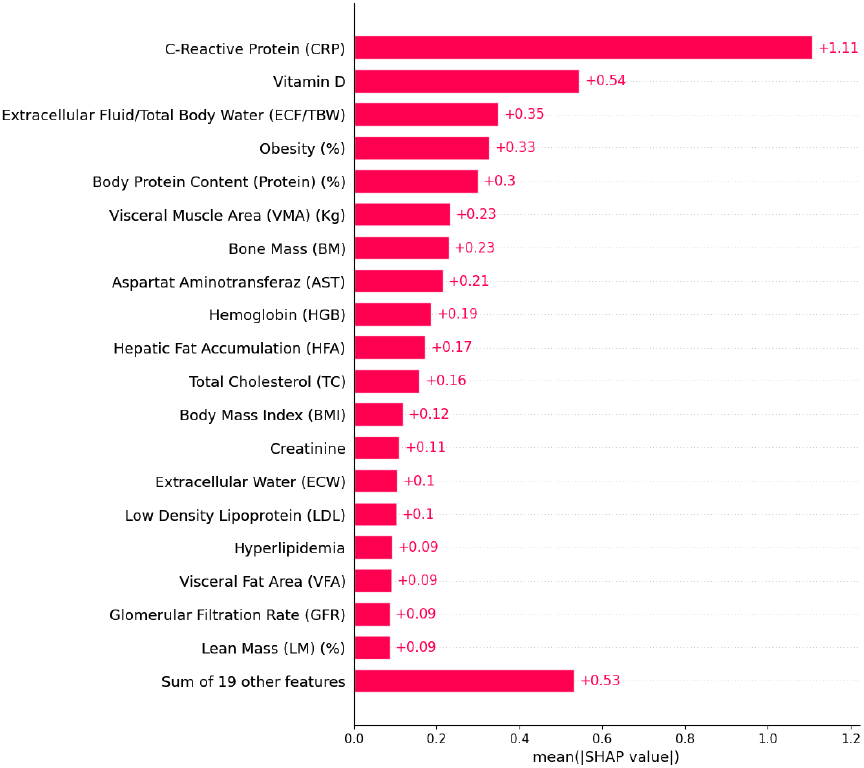
Global feature importance ranked by mean absolute SHAP value.

Also, local SHAP values provide patient-specific explanations. Table III shows the top ten contributing features for a representative patient, revealing individualized metabolic and inflammatory risk factors.

**TABLE 3:** Top 10 LOCAL SHAP ATTRIBUTIONS FOR A REPRESENTATIVE PATIENT.

| Feature | Value | SHAP |
| --- | --- | --- |
| C-Reactive Protein (CRP) | 2.00 | 0.8216 |
| Creatinine | 0.61 | -0.3794 |
| Hepatic Fat Accumulation (HFA) | 2.00 | 0.2428 |
| Vitamin D | 20.20 | 0.2165 |
| Body Protein Content (%) | 16.15 | -0.1797 |
| Obesity (%) | 29.76 | -0.1727 |
| Intracellular Water (ICW) | 29.00 | 0.1365 |
| Body Mass Index (BMI) | 33.10 | 0.1273 |
| Extracellular Water (ECW) | 20.00 | -0.1156 |
| Bone Mass (BM) | 3.40 | -0.0957 |

## V. Discussion

The experimental results show how our proposed pipeline produces accurate and physiologically meaningful predictions while at the same time maintaining interpretability. The twin-constrained classifier achieves an accuracy of 0.81 and a macro–F1 of 0.81 (Table I), with an AUROC of 0.85 indicating reliable discrimination between gallstone and non-gallstone patients.

Global explainability analysis aligns with known clinical patterns. The SHAP summary diagram in Fig. 3 emphasizes C-Reactive Protein (CRP), Vitamin D, the extracellular fluid ratio (ECF/TBW), and obesity as the most significant factors influencing the predictions. These markers serve as validated indicators of inflammation and metabolic disturbance, underscoring that the model depends on physiologically credible signals. Additional contributors, including body protein content, visceral muscle area, and hepatic fat accumulation, represent significant pathways related to metabolic and hepatic risk, in accordance with existing literature. The international rankings depicted in Table II further corroborate these patterns.

**Fig. 3.**
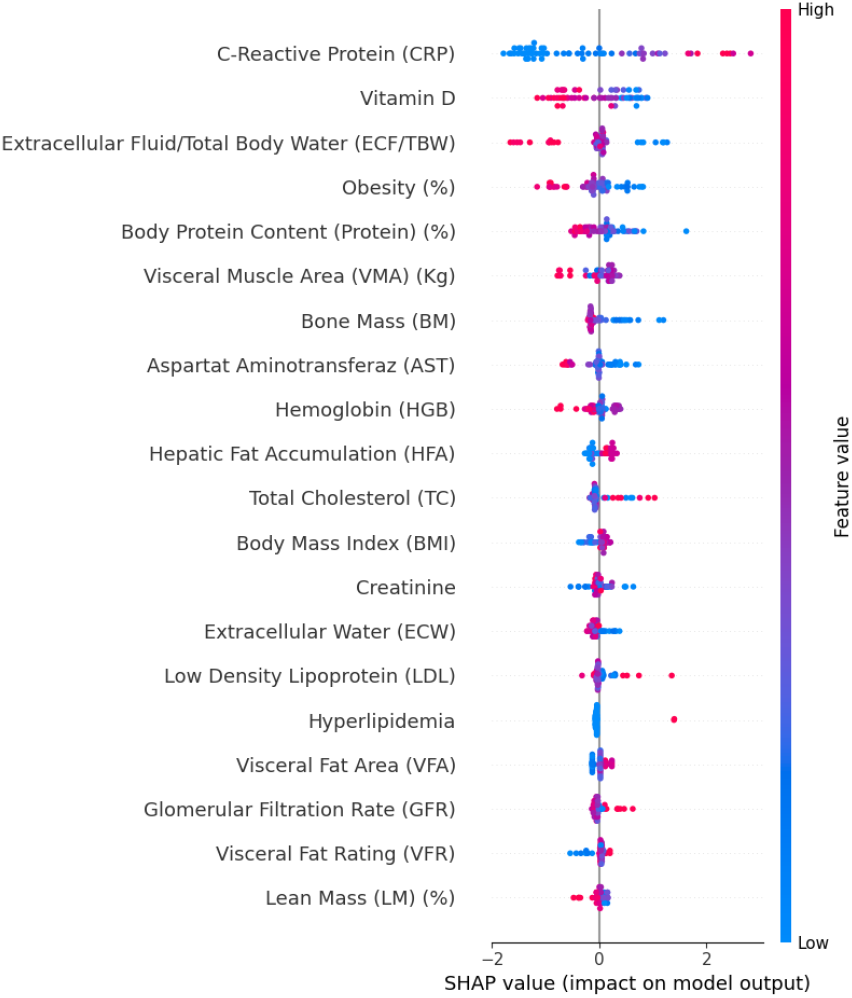
SHAP summary plot showing global feature contributions to the model.

On the other hand,local explanations offer personalized insights. Table III presents a summary of the most significant SHAP features for a representative patient. Elevated CRP levels, increased intracellular water content, elevated BMI, and low Vitamin D status emerge as primary risk factors. Such patient-specific explanations demonstrate how the model customizes global patterns to an individual’s physiological profile, a crucial capability for personalized decision support.

A fundamental result of the framework’s design is that the digital duplicate restricts the internal representation of the classifier. This reduces spurious correlations and ensures that predictions remain consistent with simulated patient trajectories. In practice, this enhances the credibility of explanations and stabilizes SHAP attributions, as demonstrated by the uniform feature-value gradients depicted in Fig. 3. The twin also assesses counterfactual recommendations by excluding biologically implausible modifications.

## VI. XAI Mechanisms and Techniques

PA-XDT uses several forms of explanation to make its predictions easier to understand and to show how each result was produced.

### A. Feature Attribution and Sensitivity Analysis

The system uses SHAP to show which features matter most. For each patient, it highlights the factors that push the prediction up or down, and at the population level it summarizes the features that carry the most weight overall. Small changes to the inputs are also tested to see which variables the model is most sensitive to and how they interact.

### B. Counterfactual Explanations and What-If Scenarios

Counterfactuals suggest small, practical changes that could move a patient’s predicted outcome in a different direction. The digital twin checks each suggestion and keeps only those that make biological sense. This allows users to explore simple ‘what-if’ situations and compare which changes are most realistic and helpful.

### C. Twin-Guided Explainability

The digital twin helps keep explanations grounded. If an attribution or counterfactual does not agree with how a patient’s physiology is expected to behave, it is flagged for user review. This ensures that the explanations reflect patterns that could occur in practice.

### D. Feedback Integration and Adaptive Learning

Feedback from clinicians and patients can be used to adjust and improve the explanations. If concerns arise, the system reviews the model’s behavior. If the feedback supports the explanation, it strengthens confidence in those patterns. Over time, this process helps keep the explanations clear and clinically aligned.

## VII. Use-Case Scenarios

The PA-XDT framework is designed for personalized health and wellness management, supporting a variety of real-world applications where interpretable predictions and patient-specific simulations are critical.

### A. Chronic Disease Management

Patients with chronic conditions such as hypertension, diabetes, or cardiovascular disease can benefit from continuous monitoring through wearable devices and home sensors. The digital twin simulates the patient’s physiological responses to lifestyle changes, medications, or interventions, while the XAI module provides interpretable explanations of predicted risks. Clinicians can evaluate counterfactual scenarios, such as how dietary adjustments or greater physical activity influence health, leading to tailored treatment recommendations.

### B. Lifestyle and Wellness Optimization

Healthy individuals or patients at risk can use PA-XDT to optimize lifestyle choices such as sleep, exercise, nutrition, and stress management. The twin simulates how daily behaviors impact long-term health outcomes, and the XAI module provides clear guidance on which behaviors have the greatest impact. Counterfactual explanations allow users to explore minimal changes that could significantly improve outcomes, supporting proactive, data-driven wellness decisions.

### C. Patient Education and Engagement

PA-XDT empowers patients by providing understandable explanations of their health data and predicted outcomes, improving engagement and adherence. Visualizations and simplified rule-based summaries allow patients to comprehend which factors influence their health and how small changes can affect outcomes, fostering shared decision-making with clinicians.

## VIII. Conclusion

We presented PA-XDT, a provenance-aware and explainable digital twin framework for transparent, patient-centered health prediction. The system uses a lightweight LSTM to model short-term physiological transitions and to guide the interpretability pipeline. A two-phase classifier aligns its latent space with twin-derived dynamics, improving robustness and clinical coherence. Global SHAP attributions and twin-validated counterfactuals show that the model relies on meaningful physiological markers, including inflammation, metabolic function, and body composition. The provenance layer adds reproducibility by logging predictions, explanations, and counterfactual outputs.

The current implementation is limited to numerical features and short temporal windows. Future work will extend the framework to multimodal data, longer horizons, federated learning, and adaptive feedback. PA-XDT offers a practical foundation for interpretable and clinically aligned digital health decision support.

## Data Availability

A fully anonymized version of the dataset has since been made publicly available under a Creative Commons Attribution 4.0 International (CC BY 4.0) license through the UCI Machine Learning Repository.

https://archive.ics.uci.edu/dataset/1150/gallstone-1

